# Resilience, mental health, sleep, and smoking mediate pathways between lifetime stressors and Multiple Sclerosis severity

**DOI:** 10.1101/2024.02.06.24302405

**Authors:** Carri S. Polick, Hala Darwish, Leo Pestillo de Olivera, Ali Watson, Joao Ricardo Nickenig Vissoci, Patrick S. Calhoun, Robert Ploutz-Snyder, Cathleen M. Connell, Tiffany J. Braley, Sarah A. Stoddard

**Affiliations:** School of Nursing, Duke University, Durham, NC, USA; VA Healthcare System, Durham, NC, USA; Division of Multiple Sclerosis & Neuroimmunology, Department of Neurology, Michigan Medicine, Ann Arbor, MI, USA; School of Nursing, University of Michigan, Ann Arbor, MI, USA; Health Promotion Department, Cesumar University, Brazil; School of Medicine, Duke University, Durham, NC, USA; Department of Neurosurgery, Duke University, Durham, NC, USA; Department of Psychiatry, Duke University, Durham, NC, USA; School of Public Health, University of Michigan, Ann Arbor, MI USA

## Abstract

**Intro:** Lifetime stressors (e.g., poverty, violence, discrimination) have been linked to Multiple Sclerosis (MS) features; yet mechanistic pathways and relationships with cumulative disease severity remain nebulous. Further, protective factors like resilience, that may attenuate the effects of stressors on outcomes, are seldom evaluated.

**Aim:** To deconstruct pathways between lifetime stressors and cumulative severity on MS outcomes, accounting for resilience.

**Methods:** Adults with MS (N=924) participated in an online survey through the National MS Society listserv. Structural Equation Modeling was used to examine the direct and indirect effect of lifetime stressors (count/severity) on MS severity (self-reported disability, relapse burden, fatigue, pain intensity and interference), via resilience, mental health (anxiety and depression), sleep disturbance, and smoking.

**Results:** The final analytic model had excellent fit (GFI=0.998). Lifetime stressors had a direct relationship with MS severity (β=0.27, p<.001). Resilience, mental health, sleep disturbance, and smoking significantly mediated the relationship between lifetime stressors and MS severity. The total effect of mediation was significant (β=0.45).

**Conclusions:** This work provides foundational evidence to inform conceptualization of pathways by which stress could influence MS disease burden. Resilience may attenuate effects of stressors, while poor mental health, smoking, and sleep disturbances may exacerbate their impact. Parallel with usual care, these mediators could be targets for early multimodal therapies to improve disease course.

**Highlights:**

- Lifetime stressors directly and indirectly relate to MS severity.
- Resilience, mental health, sleep, and smoking were mediators.
- Resilience attenuated the effects of stressors on mental health, sleep, and smoking.
- Multi-modal interventions are needed to help improve MS symptom severity.

## 1. Background

Multiple sclerosis (MS) is a chronic immune mediated disease of the central nervous system associated with high physical, emotional, and cognitive morbidity that contribute to personal and societal costs^1^. Although immunomodulatory treatments designed to slow or prevent MS disability progression have advanced over the last decade^1^, these treatments do not adequately prevent or reverse many of the “invisible” symptoms associated with MS (e.g., fatigue, mood disturbances, pain). These invisible symptoms negatively affect both physical and cognitive wellbeing. A preventative lens focused on adjacent interventional targets to prevent additional morbidity in parallel with disease modifying therapies (DMTs) may help lessen the overall disease burden trajectory and associated costs, while improving quality of life.

Although stress has long been linked to MS, the exact mechanism and relationships remain nebulous with mixed evidence^2,3^. Since the landmark Adverse Childhood Experiences (ACEs) study^4^, overwhelming evidence suggests that childhood stress and adversity (e.g., violence, neglect) are associated with a plethora of negative health outcomes including many of the leading causes of mortality^5^. The relationship between stress and negative health outcomes is thought to be mediated in part by increased inflammatory processes and risky health behaviors (e.g., substance use)^4,5^.

Recently, there has been growing interest in the association between childhood stress and immune mediated diseases including MS.^6,7^ While the literature on this topic is still scarce, preliminary evidence suggests that childhood stress is associated with decreased health-related quality of life and increased fatigue in people with MS (PwMS)^8,9^. The literature examining the relationship between childhood stress and adversity with MS related outcomes is currently limited by a failure to include a lifetime approach to the assessment of stressful life events^10^. The childhood stress experience impacts the adult stress experience (e.g., increased perception of stress, pain), and failure to account for both could limit insight on this topic^11,12^. To date, there has been only one published study to apply a comprehensive lifespan approach to examine the association between stress and MS features. Results indicated that both child and adult stressors were related to worse disability in PwMS^13^. Yet, when evaluating relapse burden, childhood stress was not a significant predictor after adult stressors were added to the model, indicating shared contributions and the possibility that proximal stressors may have a greater impact on health outcomes. Results highlight the importance of including stressors across the lifetime when examining contributions of stressors on MS related outcomes^13^.

### 1.1 Potential mediators

There has also been little research examining the relationship between stressful events starting in childhood and MS severity that have accounted for other variables known to be related to both stress and MS disease burden. The association between lifetime stressors and MS health is likely not only direct. Indeed, previous studies have identified potential mediators, which are variables that are caused by the independent variable and influence the dependent variable. For example, it is well known that stress exposure is associated with worse mental health, health risk behaviors (e.g., tobacco smoking), and increased sleep disturbance^12,14,15^. In turn, psychological distress, smoking, and sleep problems are known to be related worse MS outcomes^16-18^. PwMS have a higher lifetime prevalence of mental health challenges including depression, anxiety, bipolar disorder, adjustment disorders, and psychotic disorders^19-21^. The prevalence and severity of sleep disturbances, including poor sleep quality, sleep fragmentation, sleep apnea, insomnia, and restless legs syndrome (RLS), are increased in PwMS^22-24^. Despite their high prevalence and impact, sleep disturbances often go unrecognized and untreated^25^, contributing to other symptoms as well as diminishing quality of life in PwMS^24,26,27^. Lastly, tobacco smoking is particularly harmful in the context of MS^28^. Through inflammatory and immune processes, smokers have 50% higher risk of developing MS, progress faster through the disease continuum with increased disability and relapses^29-31^. Further, some evidence linked smoking to reduced effectiveness of some DMTs^32,33^, yet disability and the overall disease course can improve after cessation^30^. Smoking has rarely been accounted for while evaluating relationships between childhood stress and MS, and smoking, mental health issues, and sleep have never been evaluated as mediators between lifetime stress and MS outcomes^10^.

Another important potential mediator of the association between stressful life event and health outcomes is psychological resilience. Resilience is defined as “the process and outcome of successfully adapting to difficult or challenging life experiences, especially through mental, emotional, and behavioral flexibility and adjustment to external and internal demands”^34^. Individuals with MS benefit from resilience as it empowers them to surmount challenges and barriers associated with the uncertainties of MS and its symptoms^35^. PwMS with heightened resiliency appear to have enhanced quality of life and better mental health^36,37^. Numerous determinants of resilience in MS have been identified. Individual characteristics, such as personality traits like optimism and problem-focused coping strategies^38,39^. Additionally, social support has consistently emerged as a critical factor in promoting resilience and improving health-related quality of life for individuals with MS^37^. Yet, resilience has not yet been evaluated as a potential mediator between lifetime stressors and health outcomes amongst PwMS.

### 1.2 Aim

The aim of this study was to examine the direct and potential mediating pathways between lifetime stressors and severity of MS symptoms through other factors known to be associated with both exposure to stressors and MS symptoms. More specifically, we examined resilience, mental health, smoking, and sleep disturbances as mediators between lifetime stressors and MS severity.

## 2. Methods

This study is a secondary data analysis of the Stress-MS dataset approved by Duke University IRB. PwMS were recruited to complete an online survey through the US National MS Society email listserv in October 2021. Additional details can be found in the original papers^13,40^. Eligible participants were US-based adults with MS who consented to the online survey during the month of recruitment. Those without a diagnosis of MS were ineligible. The described methodological procedures followed the indications of the APA Standards for Quantitative Research^41^ and the checklist for Structural Equation Modeling described by Kang and Ahn^42^.

### 2.1 Measures and model indicators

#### 2.1.1 Lifetime stressors

Lifetime Stressors included a count of stressors and cumulative sum severity of stressors, measured by the Stress and Adversity Inventory - STRAIN^43^. The STRAIN tool is a NIMH/RDoC (Research Domain Criteria) recommended instrument, which effectively and dependably evaluates an individual’s aggregated experience with stressors spanning childhood and adulthood, totaling 55 different stressors. Examples include childhood abuse, financial strain, inter-personal or community violence, and discrimination. Stressor severity items are scored on a scale from “Very slightly or not at all” to “Extremely” and then summed. Higher scores for severity or count represent higher stress levels. Stressors were assessed as the cumulative adult count/severity.

#### 2.1.2 Resilience

The Multiple Sclerosis Resiliency Scale (MSRS)^44^ is a comprehensive self-report measure specifically tailored to the unique challenges and experiences faced by individuals living with MS and spans from before their disease to the present. The MSRS contains 25 items on a 4-point Likert scale (1 = “strongly disagree” to 4 = “strongly agree”), the higher the score, the greater the level of resilience. The measure includes five subscales: Emotional and Cognitive Strategies (13 items; e.g. “I can deal with the stress related to my MS”); Physical Activity and Diet (3 items, e.g., “Exercising helps me reduce my stress”); MS Peer Support (2 Items; e.g. “I have learned to reach out to others with MS”); Support from Family and Friends (5 Items; e.g. “I have supportive relationships on while I can rely”); and Spirituality (2 Items, e.g. “Having belief in a higher power helps me deal with my MS”). The MSRS total score is calculated by adding the 25 items together, this total score was used in the model. The validity and reliability study showed good internal consistency results for the total score (Cronbach’s α = 0.84) and ranged from fair to excellent for the five subscales (0.74 to 0.91). Since some MSRS questions ask about a longer length of time retrospectively than the other measures, it was modeled as the first mediator.

#### 2.1.3 Mental Health Issues

Mental Health Issues was comprised of two Patient Reported Outcomes Measurement Information System (PROMIS) tools^45,46^. PROMIS Anxiety (4 item) and PROMIS Depression (4 item) captured symptoms of both. Items are rated on a 5-point Likert scale, with respective scores ranging from 4 to 20. Elevated scores on these scales are indicative of more severe symptomatology.

#### 2.1.4 Sleep Disturbance

Sleep disturbance was measured with the PROMIS Sleep^46^ (4 item) which captures sleep quality and sleep difficulties over the past week on a 5-point Likert scale. Examples include “My sleep quality was…” (very good to very poor), and “I had a problem with my sleep” (Not at all to very much). A sleep disturbance score is obtained by the sum of the 4 items responses.

#### 2.1.5 Smoking

The variable Smoking is assessed with a single item self-report history with tobacco (Never smoker; Ex-smoker; Current or social smoker).

#### 2.1.6 MS severity

MS severity was assessed through fatigue, pain intensity, pain interference, disability level, and relapse burden changes since Covid-19 onset.

Fatigue over the past week was measured by PROMIS Fatigue-MS (8 item), which was developed from the PROMIS fatigue item bank with the aid of input from MS patients and clinical experts^47^. This scale has a 5-point Likert scale from “Never” to “Always”. Examples include “How often are you too tired to think clearly?” and “How often did you have trouble finishing things because of your fatigue?” The scoring of the PROMIS Fatigue-MS 8 item is based on a T-score metric, whereby elevated scores are indicative of heightened levels of fatigue.

Pain intensity over the past week was measured by PROMIS Pain Intensity^46^ (3 item), with a 5-point Likert scale from “Had no pain” to “Very severe pain”. Examples include “How intense was your average pain?” and. “How intense was your pain at its worst?”

Pain Interference over the past week was measured by PROMIS Pain Interference^46^ (8 item) with a 5-point Likert scale from Not at all to Very much. Examples include “How much did pain interfere with your day-to-day activity?” and “How much did pain interfere with things you usually do for fun?”

Relapse burden was measured by participants reporting how their relapses have changed since COVID-19. For example, a worse burden (e.g., more frequent, disabling, painful, longer relapses), no change, a lighter burden (e.g., less painful, fatiguing, shorter), or no relapses between March 2020 and data collection in October 2021.

Disability was measured by the Patient Determined Disease Steps (PDDS), an accepted measure for measuring MS disability. Patient Determined Disease Steps (PDDS) is a validated patient reported scale of MS disability^48,49^. This is a 1-item scale with scores ranging from 0-8 representing the progression from normal function (0) to being bedridden (8). Scores are commonly converted to categorical outcomes for interpretability of mild, moderate, or severe disability^48^, which was implemented in this study.

### 2.2 Measurement and Structural Model setting

We first developed a measurement model that assessed the structure and adequacy of the measurement of the latent variables. In this model, we had three latent variables, defined in the measure session (Lifetime Stressors, Mental Health Issues, and MS Severity). Lifetime Stressor is an exogenous variable and the main predictor in our model. Mental Health is a mediator, functioning as endogenous and exogenous. MS severity is an endogenous variable, the main outcome in the model.

Next, we developed a causal diagram based on previous literature to display the hypothesized association between Lifetime Stressors and MS severity, mediated by mental health severity, sleep disturbance and smoking. The structural model depicts the hypothesized association paths between the exogenous variables with MS severity, mediated by mental health issues, sleep disturbance and smoke. Our hypothesized model assumed a path from Lifetime Stressors to Resilience (mediation) and a direct path to MS severity. Resilience has a path to Mental Health Issues, Sleep Disturbance and Smoking (mediation) and a direct path to MS Severity. Mental Health Issues, Sleep Disturbance and Smoking have a path to the outcome, MS severity.

The model was specified to be identified following the consideration that a SEM model must have fewer parameters than observed variables. According to Figure 1, a difference between the number of observed variables included in the model (45) and the number of parameters to be estimated (31) results in 14 degrees of freedom of the mode.

**Figure 1.**
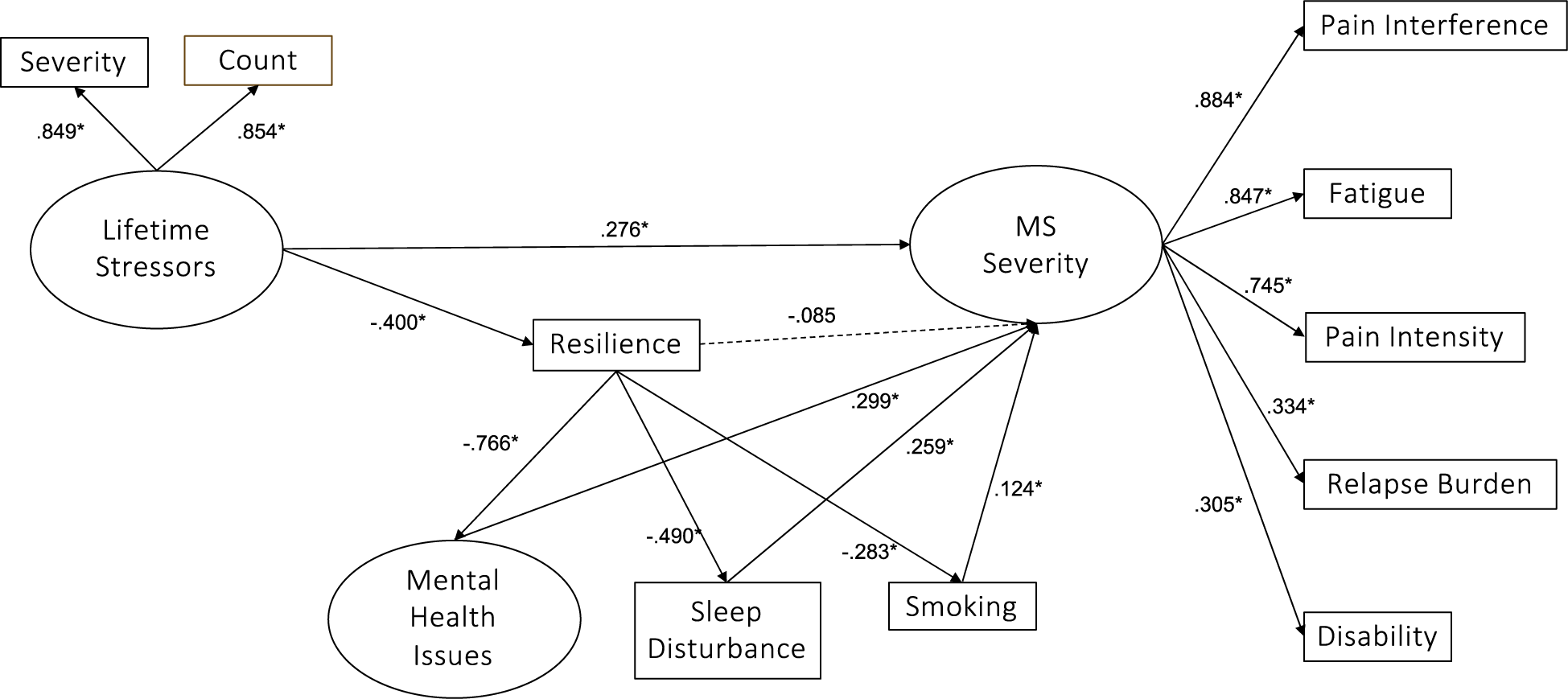
Measurement model: impact of Lifetime Stressors on MS Severity mediated by Resilience, Mental Health Issues, Sleep Disturbance and Smoking.

### 2.3 Data analysis

Summary statistics were generated for the participant demographics as well as for the measures employed in the model. We used mean results (with confidence interval), standard deviation, frequency, and percentage for sociodemographic variables. Polychoric correlations were used to depict the association of all indicators included in the. We also used Structural equation modeling (SEM)^50^ approach to evaluate the association of Stressors and Multiple Sclerosis Severity, and the mediation role of resilience, mental health, sleep disturbances and smoking. All analyses were undertaken using a significance level of p < 0.05. and conducted with R Language for Statistical Computing, version 4.3.0^51^, Correlation structures were estimated using the qgraph package^52^ and SEM models were fit using the lavaan package version 0.6-16^53^.

#### 2.3.1 SEM estimation, evaluation and modification

We estimated the SEM model using Weighted Least Squares Mean and Variance Adjusted (WLSMV) method. This approach has been reported to adjust for mixed-data models including categorical data indicators. This is a robust estimator that is less sensitive to violations of normality than other estimators maximum likelihood based estimators^50^. For the treatment of missing data, we used an imputation of values for variables from Likert scales, and the method of most frequent category for categorical variables^54^.

We iteratively tested the SEM models and presented measures of association for the direct, indirect, and total coefficients of association between variables. We examined the goodness of fit of the constructed models using the chi-squared (χ2) test with the degree of freedom to compare the proposed model to a saturated model. To complement the result of the chi-square test, we used other measures, GFI, AGFI, RMSEA (with CI), CFI, and TLI. The GFI (Goodness-of-Fit Index) and AGFI (Adjusted GFI) indices are indices of general adjustment of the model, and their values close to 1 are considered ideal, with values above 0.90 considered acceptable. The CFI (Comparative Fit Index) and TLI (Tucker Lewis Index) measures are indices that compare the performance of the model with the null model (without variables), for these measures, the ideal values are close to 1, being at least 0.90 an optimal value. The RMSEA (Root Mean Square Error of Approximation) is a parsimony index that attempts to correct flaws in the chi-square measure, considering ideal values below 0.08^55,56^.

We tested the hypothesized model and inspected the modification indices to evaluate the model structure changes, evaluating the relationship of the errors of measurement and endogenous variables. We also tested variations of the model to evaluate its plausibility, equivalency and parsimony with a fully mediated path and partially mediated path.

## 3. Results

Aligning with much MS research, participants were mostly women (83.7%), with average age of 48.7 (SD 12.8, ranging from 18 to 85 years). The mean age of onset of symptoms was 30.4 years (SD 10.4), and the mean age at diagnosis was 35.8 years (SD 10.3). The predominant type of MS was Relapsing Remitting (79.1%), followed by Progressive Relapsing (1.4%). Regarding education, 36.6% of the participants had a bachelor’s degree and 26.8% had a master’s degree. Household income varied widely, with the most common income range being over $150,000 (20.9%), followed by $50,000 to $74,999 (16.1%) and $75,000 to $99,999 (14.9%) (Table 1). Correlations among study variables are found in Table 2.

**Table 1.**
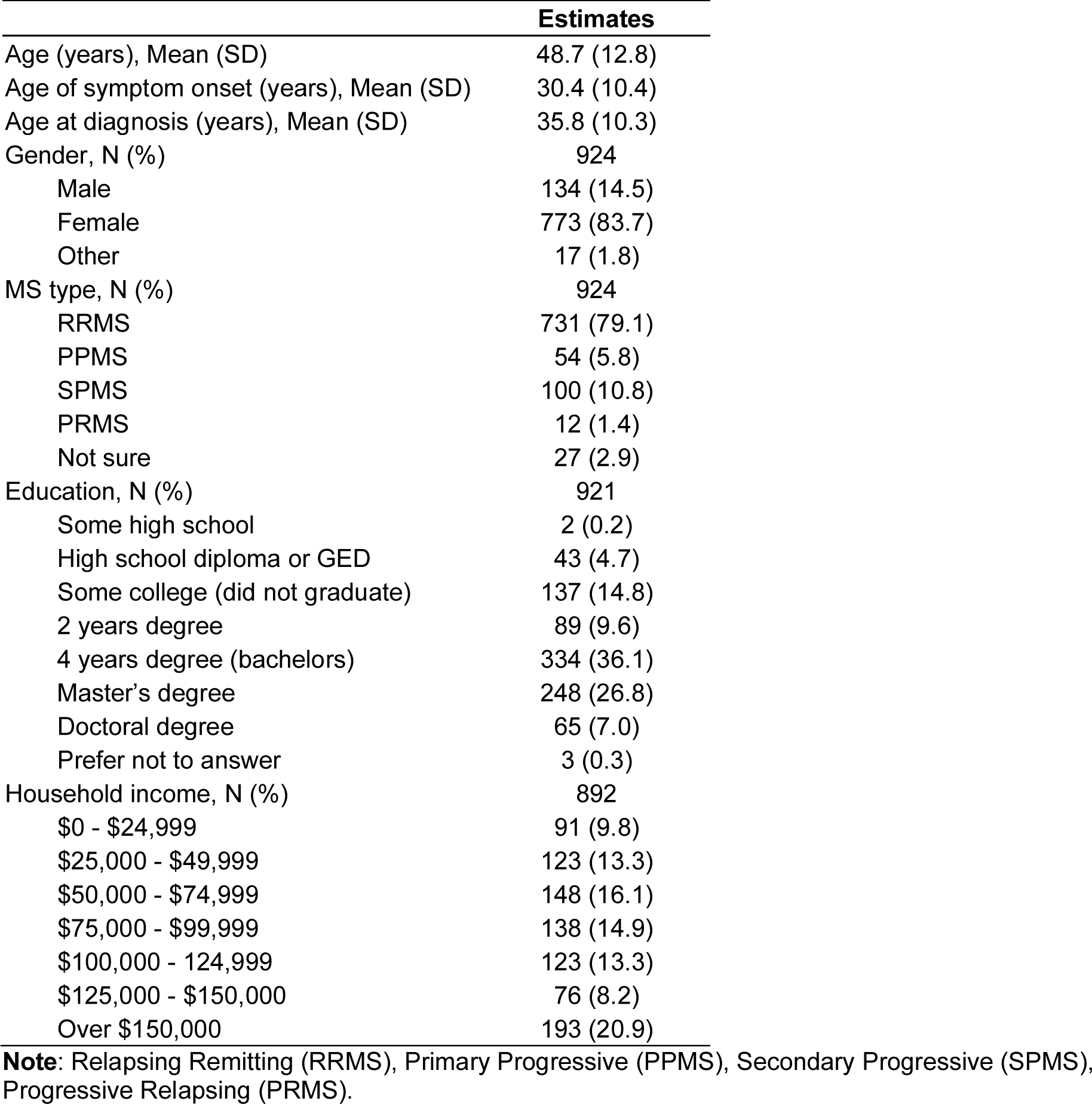
Subject demographics.

|  | **Estimates** |
| --- | --- |
| Age (years), Mean (SD) | 48.7 (12.8) |
| Age of symptom onset (years), Mean (SD) | 30.4 (10.4) |
| Age at diagnosis (years), Mean (SD) | 35.8 (10.3) |
| Gender, N (%) | 924 |
| Male | 134 (14.5) |
| Female | 773 (83.7) |
| Other | 17 (1.8) |
| MS type, N (%) | 924 |
| RRMS | 731 (79.1) |
| PPMS | 54 (5.8) |
| SPMS | 100 (10.8) |
| PRMS | 12 (1.4) |
| Not sure | 27 (2.9) |
| Education, N (%) | 921 |
| Some high school | 2 (0.2) |
| High school diploma or GED | 43 (4.7) |
| Some college (did not graduate) | 137 (14.8) |
| 2 years degree | 89 (9.6) |
| 4 years degree (bachelors) | 334 (36.1) |
| Master's degree | 248 (26.8) |
| Doctoral degree | 65 (7.0) |
| Prefer not to answer | 3 (0.3) |
| Household income, N (%) | 892 |
| \$0 - \$24,999 | 91 (9.8) |
| \$25,000 - \$49,999 | 123 (13.3) |
| \$50,000 - \$74,999 | 148 (16.1) |
| \$75,000 - \$99,999 | 138 (14.9) |
| \$100,000 - 124,999 | 123 (13.3) |
| \$125,000 - \$150,000 | 76 (8.2) |
| Over \$150,000 | 193 (20.9) |

**Note:** Relapsing Remitting (RRMS), Primary Progressive (PPMS), Secondary Progressive (SPMS), Progressive Relapsing (PRMS).

**Table 2:** Correlation between model variables.

| Variable | 1 | 2 | 3 | 4 | 5 | 6 | 7 | 8 | 9 | 10 | 11 |
| --- | --- | --- | --- | --- | --- | --- | --- | --- | --- | --- | --- |
| 1. Resilience | — |  |  |  |  |  |  |  |  |  |  |
| 2. Stress Count | -0.228*** | — |  |  |  |  |  |  |  |  |  |
| 3. Stress Severity | -0.231*** | 0.725*** | — |  |  |  |  |  |  |  |  |
| 4. Mental Health Severity | -0.645*** | 0.299*** | 0.300*** | — |  |  |  |  |  |  |  |
| 5. Sleep Disorder | -0.364*** | 0.221*** | 0.230*** | 0.429*** | — |  |  |  |  |  |  |
| 6. Smoke | -0.181*** | 0.224*** | 0.203*** | 0.133*** | 0.105** | — |  |  |  |  |  |
| 7. Relapse Burden | -0.197*** | 0.098** | 0.092** | 0.254*** | 0.209*** | 0.047 | — |  |  |  |  |
| 8. Disability | -0.190*** | 0.156*** | 0.082* | 0.099** | 0.063 | 0.078* | -0.064 | — |  |  |  |
| 9. Pain Intensity | -0.369*** | 0.312*** | 0.309*** | 0.394*** | 0.354*** | 0.195*** | 0.238*** | 0.385*** | — |  |  |
| 10. Pain Interference | -0.450*** | 0.344*** | 0.364*** | 0.492*** | 0.402*** | 0.196*** | 0.271*** | 0.350*** | 0.839*** | — |  |
| 11. Fatigue | -0.487*** | 0.340*** | 0.333*** | 0.547*** | 0.469*** | 0.212*** | 0.281*** | 0.284*** | 0.561*** | 0.657*** | — |

Note: \* $p < .05$ , \*\* $p < .01$ , \*\*\* $p < .001$

### 4.1 SEM model estimation

Our model specification is depicted in Figure 1, along with the estimated path coefficients. This model 1 showed adequate goodness of fit indices with respect to the available data (Table 3). The latent variables in the measurement models showed adequate adjustments with factor loadings. To test the hypothesis that the effect of the lifetime stressors on the MS severity is fully mediated by the mediating variables, an alternative model (Model 2) was tested. This model presents the same characteristics as the already tested Model 1, however, without the direct paths between Lifetime Stressors and MS Severity and between Resilience and MS Severity. The fit indices for Models 1 and 2 are displayed in Table 3.

**Table 3.** Fit indices of tested Models.

| Model | $\chi^2$ | df | GFI | AGFI | RMSEA | CI | CFI | TLI |
| --- | --- | --- | --- | --- | --- | --- | --- | --- |
| Model 1:<br>with direct<br>paths | 133.445 | 37 | .998 | .996 | .053 | .044 - .063 | .976 | .964 |
| Model 2:<br>without<br>direct paths | 211.749 | 39 | .997 | .994 | .069 | .060 - .079 | .956 | .939 |

Lifetime Stressors (LFS), Mental Health Issues (MHI), Sleep Disturbance (SD), and Smoking (SMO) were all significantly and positively associated with MS severity, meaning that as stressors, mental health issues, sleep disturbance and smoking all increased, so did MS severity. Although not significant, resilience was negatively associated with MS Severity (β=0.085, P=.192), suggesting that as resilience increased the severity of MS symptoms decreased. Also notable, there was a negative association between lifetime stressors and resilience which indicates that PwMS who experienced more stressors reported lower resilience scores (Figure 1). Direct and total effect estimates are displayed in Table 4.

**Table 4.** Standardized path coefficients: Direct, indirect, and total effects.

| Model | $\beta$ | SE | p value | Confidence Interval |
| --- | --- | --- | --- | --- |
| **Direct effect** |  |  |  |  |
| LFS - RES | -0.400 | 0.027 | < 0.001 | -0.376, -0.271 |
| RES - MHI | -0.766 | 0.024 | < 0.001 | -0.554, -0.459 |
| RES - SD | -0.490 | 0.013 | < 0.001 | -0.193, -0.144 |
| RES - SMO | -0.283 | 0.004 | < 0.001 | -0.032, -0.018 |
| MHI - MSS | 0.299 | 0.015 | < 0.001 | 0.057, 0.117 |
| SD - MSS | 0.259 | 0.020 | < 0.001 | 0.105, 0.185 |
| SMO - MSS | 0.124 | 0.077 | < 0.001 | 0.119, 0.420 |
| LFS - MSS | 0.276 | 0.006 | < 0.001 | 0.032, 0.054 |
| RES - MSS | -0.008 | 0.013 | 0.192 | -0.041, 0.008 |
| **Indirect effect** |  |  |  |  |
| LFS - RES - MHI - MSS | 0.092 | 0.003 | < 0.001 | 0.009, 0.020 |
| LFS - RES - SD - MSS | 0.051 | 0.001 | < 0.001 | 0.005, 0.011 |
| LFS - RES - SMO - MSS | 0.014 | 0.001 | < 0.002 | 0.001, 0.004 |
| RES - MHI - MSS | -0.229 | 0.008 | < 0.001 | -0.061, -0.028 |
| RES - SD - MSS | -0.127 | 0.004 | < 0.001 | -0.032, -0.016 |
| RES - SMO - MSS | -0.035 | 0.002 | < 0.002 | -0.011, -0.003 |
| **Total effect** |  |  |  |  |
| LFS - MSS | 0.432 | 0.008 | < 0.001 | 0.051, 0.084 |

As seen in Table 4, lifetime stressors positively affected (i.e. worsened) MS severity indirectly through resilience and mental health issues, sleep disturbances, and smoking. In contrast to the direct pathway, the indirect effects from resilience to MS severity through mental health issues, sleep disturbance, and smoking highlight that resilience does significantly negatively relate to (i.e. reduce) MS severity through these three mediators, when not accounting for lifetime stressors. When accounting for stressors the total effect is positive, indicating that resilience may help mitigate risk but does not fully counter the effect of lifetime stressors.

## 4. Discussion

This study is the first to elucidate pathways that underlie the association between lifetime stressors and MS severity, through several addressable conditions that could ultimately serve as targets for future mitigation and prevention efforts. Research on comprehensively measured lifetime stressors and MS outcomes is scarce, and little attention has been given to positive factors such as resilience or other mediating pathways to MS severity. As expected, each exposure studied mediated relationships between stressors and adverse MS outcomes, highlighting resilience, mental health, sleep, and smoking as potential avenues for mitigating risk for more severe MS symptoms.

Surprisingly, the direct relationship from resilience to MS severity was not statistically significant, however, the indirect pathways from resilience through smoking, mental health, and sleep were significant. This suggests that resilience could be an upstream target for intervention to have earlier influence over health outcomes and/or behaviors that subsequently have downstream effects on the severity of MS disease burden. However, the direct relationship from stressors to MS severity was significant, supporting that stressors would be a further upstream target for intervention.

Indeed, the prevention or minimization of stress exposure for all people with MS requires action at the policy and public health level. On a macroscopic level, addressing financial strain, unstable housing, discrimination, and provisions for mental health support, substance use and domestic violence could help lessen acute and chronic stress and subsequently optimize health for PwMS and beyond. Recently, for the first time, the World Health Organization (WHO) listed MS disease modifying therapies on the Essential Medication List^57^. Policies that follow this lead to continue to improve MS treatment access, coverage, and affordability, may have long term impact on the financial stress of PwMS, as MS is the second most costly chronic disease in the US^1^. However, until such policies can be implemented, more needs to be done to enhance resilience and mitigate said pathways in individuals.

On an individual level, in line with our findings, Novak and Lev-Ari (2023) found a relationship between resilience, adult stress, and sleep quality in PwMS, and that higher levels of resilience are associated with better mental and physical health outcomes in the context of MS^58^. Moreover, fostering resilience is associated with improved management of challenges in aging PwMS. Higher levels of resilience contribute to better quality of life and overall well-being in older PwMS^59^.

Resilience is a multifaceted process influenced by various interrelated systems that requires a comprehensive, culturally sensitive approach when supporting individuals in stressed environments. The intricate interplay of various factors from diverse systems, such as personal, familial, social, cultural, and environmental collectively contribute to an individual’s ability to effectively cope with adversity^60,61^. Ungar and Theron’s foundational article in The Lancet Psychiatry (2020) established a theoretical framework of multisystemic resilience^62^. It underscores the significance of considering multiple factors simultaneously to understand how resilience is fostered in young individuals facing challenges^62^. This is of high significance for PwMS. The individual strengths, familial support, community resources, and cultural values will play a significant and crucial role in promoting MS recovery and positive mental health outcomes. Our results emphasize the need for more mixed-methods and multi-modal approaches and cross-cultural comparisons to capture the complexities and adaptability of resilience across diverse environments in MS^63^.

Our findings along with other research generally highlight the need for more complex, perhaps multi-modal, intervention approaches that could target resilience and additional mediators like sleep, mental health, and smoking in PwMS. Emerging research that investigated the effectiveness of a group resilience intervention for PwMS delivered through frontline services found it to be a potential way to improve psychological well-being and coping skills^64^. This group-based intervention provided a supportive environment for sharing experiences and developing effective strategies to manage the impact of the disease. In addition to resilience, emotional competencies in MS were explored by Sadeghi-Bahmani and colleagues^65,66^. Their research delves into understanding the role of emotional competencies in PwMS, with a focus on emotional regulation and processing, and has even used a multi-modal approach of mindfulness based stress reduction coupled with acceptance and commitment therapy^66^. Emotional competencies, such as emotional regulation, have been found to be associated with better psychological well-being and coping abilities in PwMS. Understanding and promoting resilience, along with emotional competencies, in PwMS can have profound implications for enhancing their ability to cope with stressors and improve health and wellness. Further research and the development of MS-focused resilience interventions which have built in components of, or are in tandem with, mental health, sleep, and smoking interventions may hold the most promising avenues for supporting PwMS to combat prevalent burdensome symptoms like fatigue.

Fatigue is highly debilitating, affects around 80% of PwMS, and is a leading cause of diminished quality of life and reduced social participation^67,68^. Despite its impact, existing interventions to treat fatigue are only modestly to moderately effective for a subset of individuals, making the identification of treatable secondary contributors a high priority. While the biological underpinnings of fatigue in MS are heterogeneous, substantial evidence has linked sleep disturbances to MS fatigue^23,69,70^. Furthermore, recent work suggest that sleep behaviors have the potential to affect response to fatigue interventions^71,72^.

Our findings that smoking is a mediator for MS severity highlights how multi-modal interventions that address resilience factors in addition to smoking cessation treatment may have the highest potential for impact. Ongoing clinical trials have shown promising results using a multi-modal approach combining 1) Cognitive Behavioral Therapy (CBT) with health education, stress, and coping components, 2) Nicotine Replacement Therapy (NRT) and 3) Transcranial Magnetic Stimulation (TMS) as a tailored smoking cessation intervention for veterans with PTSD^73^. However, there has not yet been a single- or multi-modal tailored intervention developed for MS despite evidence of disease specific concerns regarding quitting^28,74^. For example, people with MS have reported concern over the stress of quitting causing MS relapses, increased anxiety, uncertainty about adverse drug reactions, and the desire to have MS-focused information as motivation to quit^74,75^. Evidence from Australia highlights that PwMS are not satisfied with the cessation care they get, yet, we currently lack a US perspective regarding programmatic preferences and barriers for cessation treatment^75^.

### 4.1 Clinical implications

Referrals for sleep disturbances, smoking cessation clinics, and mental health treatment could be implemented in parallel with standard MS treatment to promote better health outcomes. While smoking intervention research for PwMS is lacking, there is much evidence that sleep and mental health interventions can improve the health and wellbeing of PwMS. Efforts should be made to strengthen the referral pathway from neurology and primary care providers to these adjunct treatments. An implementation science lens would be a good approach for future research to investigate how to best integrate or strengthen the screening and referral workflow to connect more PwMS to these existing services.

### 4.2 Limitations

Pertinent limitations include cross-sectional data; however, temporal ordering can at least be established with childhood and previous stressors and outcomes in adults. These findings may not generalize to all PwMS as this sample was only a small portion of the National MS Society listserv (nearly 80,000). However, this sample does generally align with conventional US MS research samples including very large studies^76^ and large studies that have similarly used the NMSS listserv^77^.

## 5. Conclusion

This work is foundational to improving conceptualization of stress, protective factors, responding behaviors, and the severity of MS disease burden. Several significant mediating pathways were found from stressors to MS severity, through resilience, sleep, mental health, and smoking. This highlights potential upstream targets for intervention across sociopolitical and individual levels. In conjunction with usual care, increasing referrals to existing treatment services for mental health, sleep disturbances, and smoking cessation could have synergistic positive benefits to the health and wellbeing of PwMS.

## Data Availability

Data in the present study are available upon reasonable request to the authors

## Notes

### Competing Interest Statement

The authors have declared no competing interest.

### Funding Statement

C.S. Polick was supported by NIH/NINR grant #T32NR016914 Complexity: Innovations for Promoting Health and Safety, the Duke Clinical and Translational Science Institute (CTSI), and the Durham VA. The content is solely the responsibility of the authors and does not necessarily represent the official views of the funders.

### Author Declarations

The IRB of Duke University gave ethical approval of this work.

